# Fragmented QRS is independently predictive of long-term adverse clinical outcomes in Asian patients hospitalized for heart failure: a retrospective cohort study

**DOI:** 10.1101/2021.07.01.21259847

**Authors:** Jeffrey Shi Kai Chan, Jiandong Zhou, Sharen Lee, Andrew Li, Martin Tan, Keith S K Leung, Kamalan Jeevaratnam, Tong Liu, Leonardo Roever, Ying Liu, Gary Tse, Qingpeng Zhang

## Abstract

**Background:** Fragmented QRS (fQRS) results from myocardial scarring and predicts cardiovascular mortality and ventricular arrhythmia (VA). We evaluated the prevalence and prognostic value of fQRS in Asian patients hospitalized for heart failure.

**Methods and results:** This was a retrospective cohort study of adult patients hospitalized for heart failure between 1^st^ January 2010 and 31^st^ December 2016 at a tertiary center in Hong Kong. The baseline ECG was analysed. QRS complexes (<120 ms) with fragmented morphology in ≥2 contiguous leads were defined as fQRS. The primary outcome was a composite of cardiovascular mortality, VA, and sudden cardiac death (SCD). The secondary outcomes were the components of the primary outcome, myocardial infarction, and new-onset atrial fibrillation. In total, 2192 patients were included, of whom 179 (8.20%) exhibited fQRS. The follow-up duration was 5.63±4.09 years. fQRS in any leads was associated with a higher risk of the primary outcome (adjusted hazard ratio (HR) 1.541 [1.185, 2.004], p=0.001), but not myocardial infarction or new-onset atrial fibrillation. fQRS in >2 contiguous leads was an independent predictor of SCD (HR 2.679 [1.252, 5.729], p=0.011). In patients without ischaemic heart disease (N=1396), fQRS in any leads remained predictive of VA and SCD (adjusted HR 3.336 [1.343, 8.282], p=0.009, and 1.928 [1.135, 3.278], p=0.015, respectively), but not cardiovascular mortality (adjusted HR 1.049 [0.662, 1.662], p=0.839).

**Conclusion:** fQRS is an independent predictor of cardiovascular mortality, VA, and SCD. Higher fQRS burden increased SCD risk. The implications of fQRS in heart failure patients without ischaemic heart disease require further studies.

## 1. Introduction

First described by Boineau and Cox in 1973, fragmented QRS (fQRS) is the manifestation of myocardial scarring on 12-lead surface electrocardiogram (ECG).(1,2) Though initially described in the context of ischaemic heart disease (IHD), fQRS has been observed in other conditions where myocardial scarring or fibrosis is present, such as hypertrophic cardiomyopathy and cardiac sarcoidosis.(2,3) The presence of fQRS has been shown to be predictive of mortality and ventricular arrhythmia (VA),(4) and several morphological criteria and classification systems have been set forth by a number of research groups.(2,5) While fQRS has been shown to be predictive of adverse cardiovascular outcomes in patients with heart failure,(4) most studies have focused on either acute outcomes of hospitalized patients, or long-term outcomes of ambulatory patients.(6,7) With the rising prevalence of heart failure in Asia,(8,9) there is an ever greater need for good prognostic markers in Asian patients with heart failure. As data on the prevalence and long-term prognostic power of fQRS in Asian patients hospitalized with heart failure are lacking, we aimed to bridge this gap in evidence with the current study.

## 2. Materials and methods

This was a retrospective cohort study approved by The Joint Chinese University of Hong Kong - New Territories East Cluster Clinical Research Ethics Committee. All patients aged 18 years old or above who were hospitalized for heart failure between 1^st^ January 2010 and 31^st^ December 2016 at a single tertiary center in Hong Kong were included. Patients who had wide QRS (≥120 ms), missing primary outcome data, and those who did not have any ECG done during the index hospitalization were excluded. The patients were identified using the Clinical Data Analysis and Reporting System (CDARS), a territory-wide database that centralizes patient information from local hospitals and ambulatory and outpatient facilities. Mortality data were obtained from the Hong Kong Death Registry, a government registry with the registered death records of all Hong Kong citizens linked to CDARS.

Patient demographics, prior comorbidities, and baseline medication usage were extracted. The baseline ECG obtained on the first heart failure admission was selected for analysis. We defined fQRS using the criteria set forth by Das et al, where fQRS was defined as the presence of an additional R wave (R’), notching in the nadir of the S wave, or presence of more than two R’ waves in at least two contiguous leads within any myocardial territory (anterior, inferior, or lateral) with QRS duration of <120 ms.(5) QRS complexes with fragmented morphology in a single lead were not classified as fQRS. The primary outcome was a composite of cardiovascular mortality, VA, and SCD. The secondary outcomes were the individual components of the primary composite outcome, myocardial infarction (MI), and new-onset atrial fibrillation (AF). All patients were followed up till 31^st^ December 2019.

All continuous variables were expressed as mean ± standard deviation and compared using Student’s t test. Dichotomous variables were compared using Fisher’s exact test. All outcomes were analyzed by univariate and multivariate Cox regression adjusting for baseline comorbidities that were significantly different between cohorts; hazard ratios (HR) were used as the summary statistic. Subgroup analyses were done to assess the prognostic power of fQRS in patients with and without ischaemic heart disease respectively, as well as the impact of fQRS burden in patients with fQRS. All p values were two-sided, and p<0.05 was considered significant. All statistical analyses were performed using SPSS software version 25.0 (IBM Corp, New York, USA). The raw data supporting the conclusions of this article will be made available on reasonable request to any of the corresponding authors, without undue reservation.

## 3. Results

In total, 2868 patients fulfilled the inclusion criteria. After excluding patients with missing ECG (N=2), missing primary outcome data (N=309), and wide QRS complex (N=373), 2182 patients were included in the analysis. We identified fQRS in 179 patients (8.20%), more of whom were males than those without fQRS (54.7% vs 46.6%, p=0.042), but were not significantly different in age (75.4±14.8 years vs 74.4±13.5 years, p=0.403). Patients with fQRS had significantly higher rates of renal diseases (p=0.011). Other baseline characteristics were not significantly different between the two cohorts (Table 1). Follow-up durations were similar (5.53±4.21 years vs 5.64±4.08 years, p=0.733). The primary composite outcome was met in 566 (25.9%) patients, while the secondary outcome of cardiovascular mortality was met in 419 (19.2%) patients, VA in 56 (2.6%) patients, SCD in 212 (9.7%) patients, MI in 353 (16.2%) patients, and new-onset AF in 829 (38.0%) patients.

**Table 1.** Baseline characteristics and follow-up durations of included patients.

|  | No fQRS<br>(N=2003) | fQRS present<br>(N=179) | p value |
| --- | --- | --- | --- |
| Follow-up duration, years | 5.64±4.08 | 5.53±4.21 | 0.733 |
| *Demographics* |  |  |  |
| Male, N (%) | 934 (46.6) | 98 (54.7) | 0.042 |
| Age, years | 74.4±13.5 | 75.4±14.8 | 0.403 |
| *Comorbidities* |  |  |  |
| Diabetes mellitus, N (%) | 616 (30.8) | 50 (27.9) | 0.447 |
| Renal diseases, N (%) | 277 (13.8) | 13 (7.26) | 0.011 |
| Hypertension, N (%) | 918 (45.8) | 69 (38.5) | 0.071 |
| Ischaemic heart disease, N (%) | 711 (35.5) | 75 (41.9) | 0.089 |
| Myocardial infarction, N (%) | 185 (9.2) | 19 (10.6) | 0.505 |
| Prior ventricular arrhythmia, N (%) | 21 (1.0) | 5 (2.8) | 0.056 |
| Prior sudden cardiac death, N (%) | 22 (1.1) | 4 (2.2) | 0.159 |
| Atrial fibrillation, N (%) | 370 (18.5) | 41 (22.9) | 0.162 |
| Stroke / TIA, N (%) | 264 (13.2) | 18 (10.1) | 0.294 |
| *Medications* |  |  |  |
| ACEI / ARB, N (%) | 1016 (50.7) | 95 (53.1) | 0.585 |
| Calcium channel blockers, N (%) | 923 (46.1) | 71 (39.7) | 0.101 |
| Beta-blockers, N (%) | 964 (48.1) | 92 (51.4) | 0.435 |
| Diuretics, N (%) | 755 (37.7) | 69 (38.5) | 0.810 |
| Nitrates, N (%) | 518 (25.8) | 57 (31.8) | 0.092 |
| Statins and fibrates, N (%) | 621 (31.0) | 51 (28.5) | 0.554 |
| Anticoagulants, N (%) | 320 (16.0) | 33 (18.4) | 0.397 |
| Antiplatelets, N (%) | 798 (39.8) | 74 (41.3) | 0.691 |

ACEI, angiotensin-converting-enzyme inhibitors. ARB, angiotensin II receptor blockers.
fQRS, fragmented QRS.

### 3.1. Prognostic value of the presence of fQRS

Cox regression (Table 2) showed that the presence of fQRS predicted the primary composite outcome in univariate analysis (HR 1.507 [1.160, 1.959], p=0.002), which remained significant after adjustments for baseline differences (HR 1.541 [1.185, 2.004], p=0.001; Figure 1). Both univariate and multivariate analyses demonstrated that fQRS was strongly predictive of cardiovascular mortality, VA, and SCD. However, there were no significant differences in the risks of MI and new-onset AF. To delineate the prognostic value of fQRS more clearly, we further adjusted the above multivariate analyses with clinical history of VA. The statistical significance of all these observations remained stable after adjustment, showing that the prognostic value of fQRS was independent of clinical history of VA.

**Table 2.**
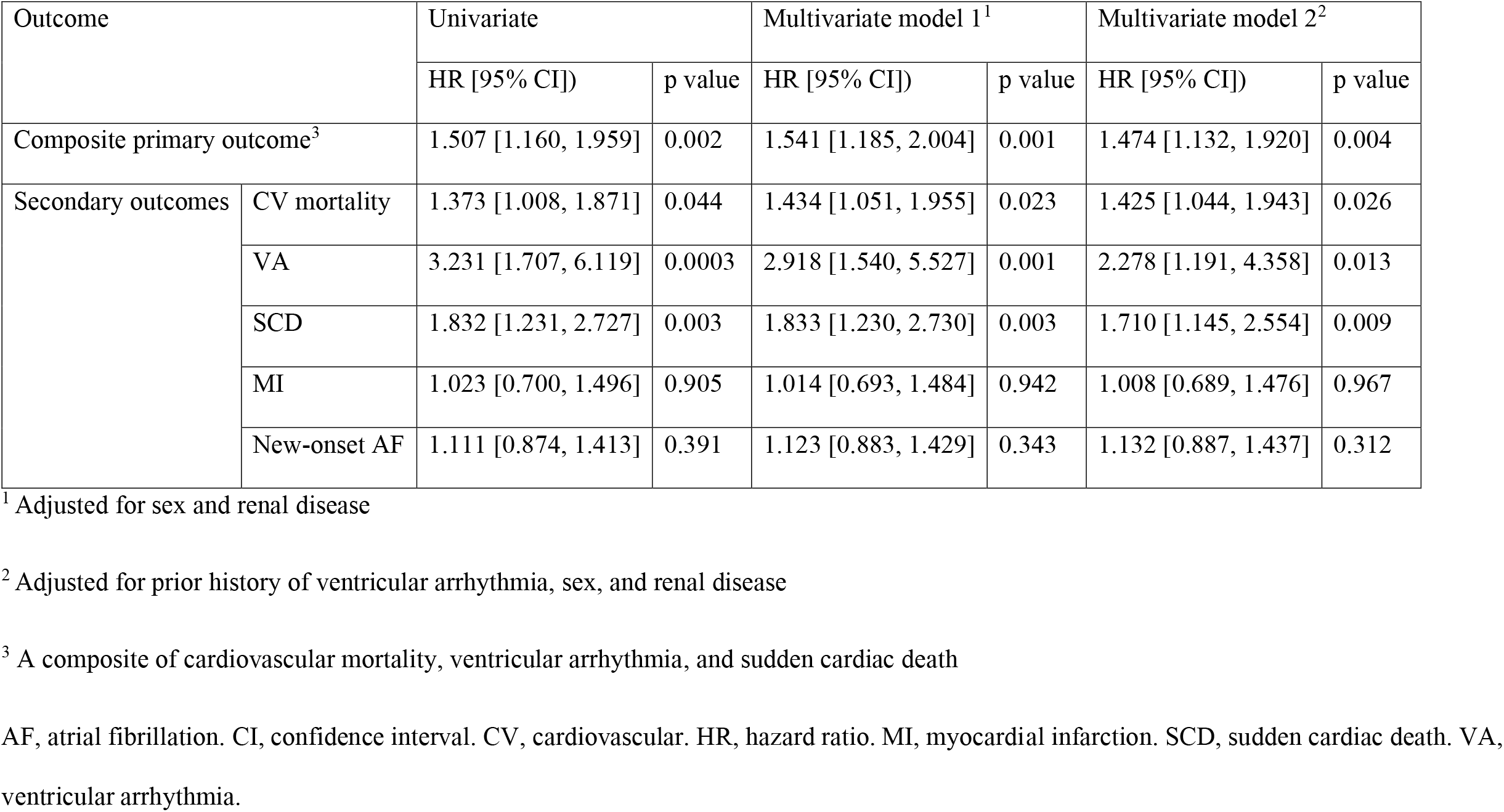
Cox regression analysis of all 2182 patients. Hazard ratios were referenced against patients without fragmented QRS.

| Outcome |  | Univariate |  | Multivariate model 1<sup>1</sup> |  | Multivariate model 2<sup>2</sup> |  |
| --- | --- | --- | --- | --- | --- | --- | --- |
|  |  | HR [95% CI]) | p value | HR [95% CI]) | p value | HR [95% CI]) | p value |
| Composite primary outcome<sup>3</sup> |  | 1.507 [1.160, 1.959] | 0.002 | 1.541 [1.185, 2.004] | 0.001 | 1.474 [1.132, 1.920] | 0.004 |
| Secondary outcomes | CV mortality | 1.373 [1.008, 1.871] | 0.044 | 1.434 [1.051, 1.955] | 0.023 | 1.425 [1.044, 1.943] | 0.026 |
|  | VA | 3.231 [1.707, 6.119] | 0.0003 | 2.918 [1.540, 5.527] | 0.001 | 2.278 [1.191, 4.358] | 0.013 |
|  | SCD | 1.832 [1.231, 2.727] | 0.003 | 1.833 [1.230, 2.730] | 0.003 | 1.710 [1.145, 2.554] | 0.009 |
|  | MI | 1.023 [0.700, 1.496] | 0.905 | 1.014 [0.693, 1.484] | 0.942 | 1.008 [0.689, 1.476] | 0.967 |
|  | New-onset AF | 1.111 [0.874, 1.413] | 0.391 | 1.123 [0.883, 1.429] | 0.343 | 1.132 [0.887, 1.437] | 0.312 |

<sup>1</sup> Adjusted for sex and renal disease
<sup>2</sup> Adjusted for prior history of ventricular arrhythmia, sex, and renal disease
<sup>3</sup> A composite of cardiovascular mortality, ventricular arrhythmia, and sudden cardiac death
AF, atrial fibrillation. CI, confidence interval. CV, cardiovascular. HR, hazard ratio. MI, myocardial infarction. SCD, sudden cardiac death. VA, ventricular arrhythmia.

**Figure 1.**
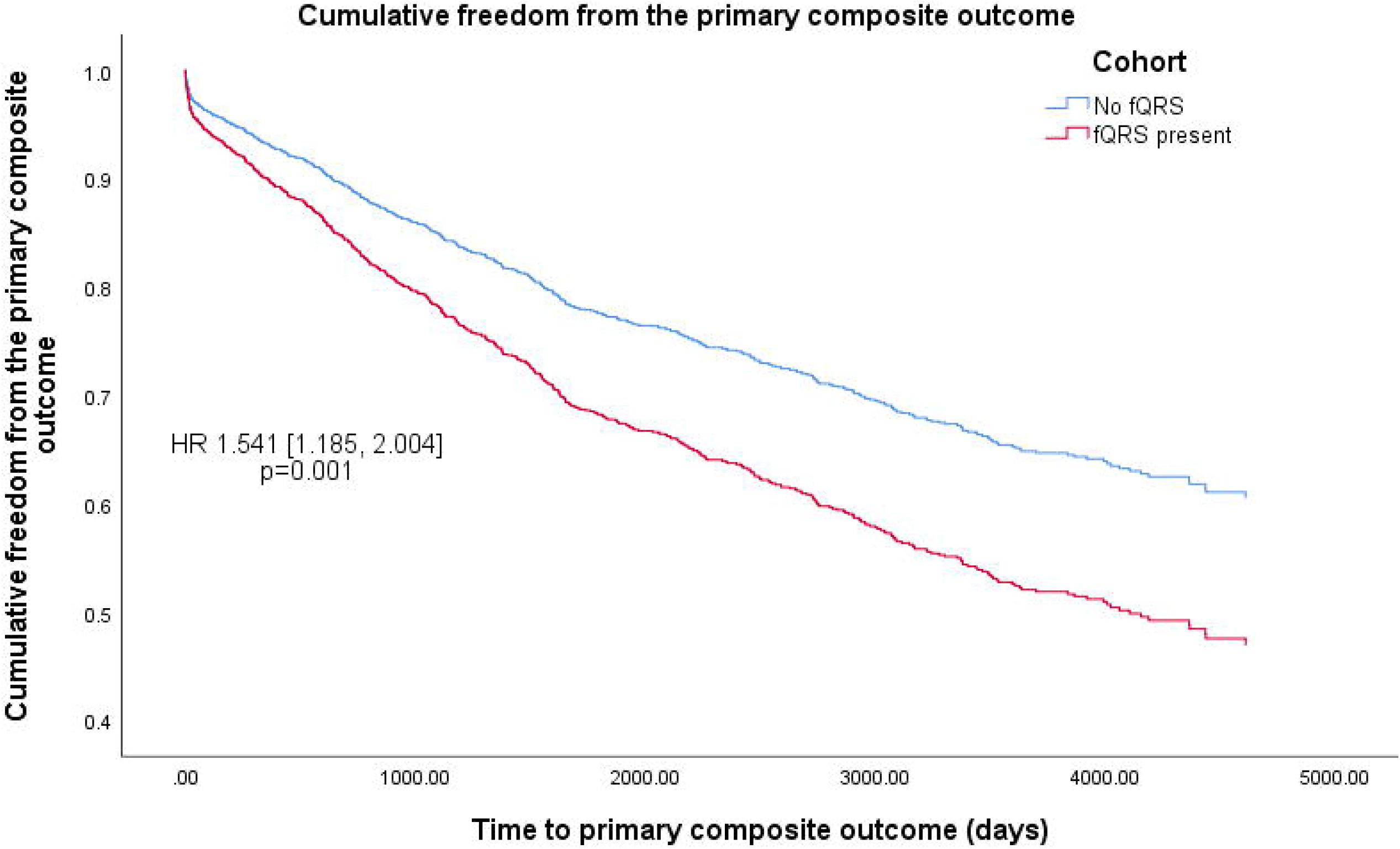
Kaplan-Meier curves of cumulative freedom from the primary composite outcome of all patients, stratified by the presence of fragmented QRS (fQRS). The hazard ratio (HR) shown was adjusted for differences in baseline characteristics.

### 3.2. Subgroup analysis by IHD status

Subgroup analysis was performed on patients with (786 (36.0%)) and without (1396 (64.0%)) IHD (Supplementary Table S1). fQRS was present in 75 (9.5%) patients with IHD, and 104 (7.4%) patients without IHD. Patients with IHD were followed up for slightly longer periods (5.82±4.13 years vs 5.29±4.00 years, p=0.003). Among patients with IHD, the primary composite outcome was met in 228 (29.0%) patients, while the outcome of cardiovascular mortality was met in 171 (21.8%) patients, VA in 28 (3.6%) patients, SCD in 83 (10.6%) patients, MI in 204 (26.0%) patients, and new-onset AF in 249 (31.7%) patients. Among patients without IHD, these were met in 338 (24.2%), 248 (17.8%), 28 (2.0%), 129 (9.2%), 149 (10.7%), and 580 (41.5%) patients, respectively. Cox regression showed that in patients with IHD, fQRS predicted the primary composite outcome (adjusted HR 1.746 [1.191, 2.559], p=0.004), cardiovascular mortality (adjusted HR 1.802 [1.173, 2.768], p=0.007), VA (adjusted HR 2.652 [1.065, 6.602], p=0.036), and SCD (adjusted HR 1.881 [1.014, 3.481], p=0.045). However, fQRS did not predict the primary composite outcome in patients without IHD (adjusted HR 1.342 [0.929, 1.938], p=0.117), despite remaining strongly predictive of VA (adjusted HR 3.336 [1.343, 8.282], p=0.009) and SCD (adjusted HR 1.928 [1.135, 3.278], p=0.015).

### 3.3. Prognostic value of fQRS burden

Patients with fQRS were stratified into those with two contiguous leads with fQRS (112 patients; 5.13% of all patients; 62.6% of patients with fQRS), and those with more than two such leads (67 patients; 3.07% of all patients; 37.4% of patients with fQRS), as summarized in Supplementary Table S2. The two subgroups were not significantly different in any baseline characteristics. Cox regression demonstrated significantly higher risk of the primary composite outcome in the latter (HR 1.841 [1.119, 3.029], p=0.016; Figure 2), driven by a significantly higher risk of SCD (HR 2.866 [1.350, 6.081], p=0.006) which remained significant even after adjustment for clinical history of VA and SCD (HR 2.679 [1.252, 5.729], p=0.011). All other outcomes were not significantly different in risk between the two subgroups.

**Figure 2.**
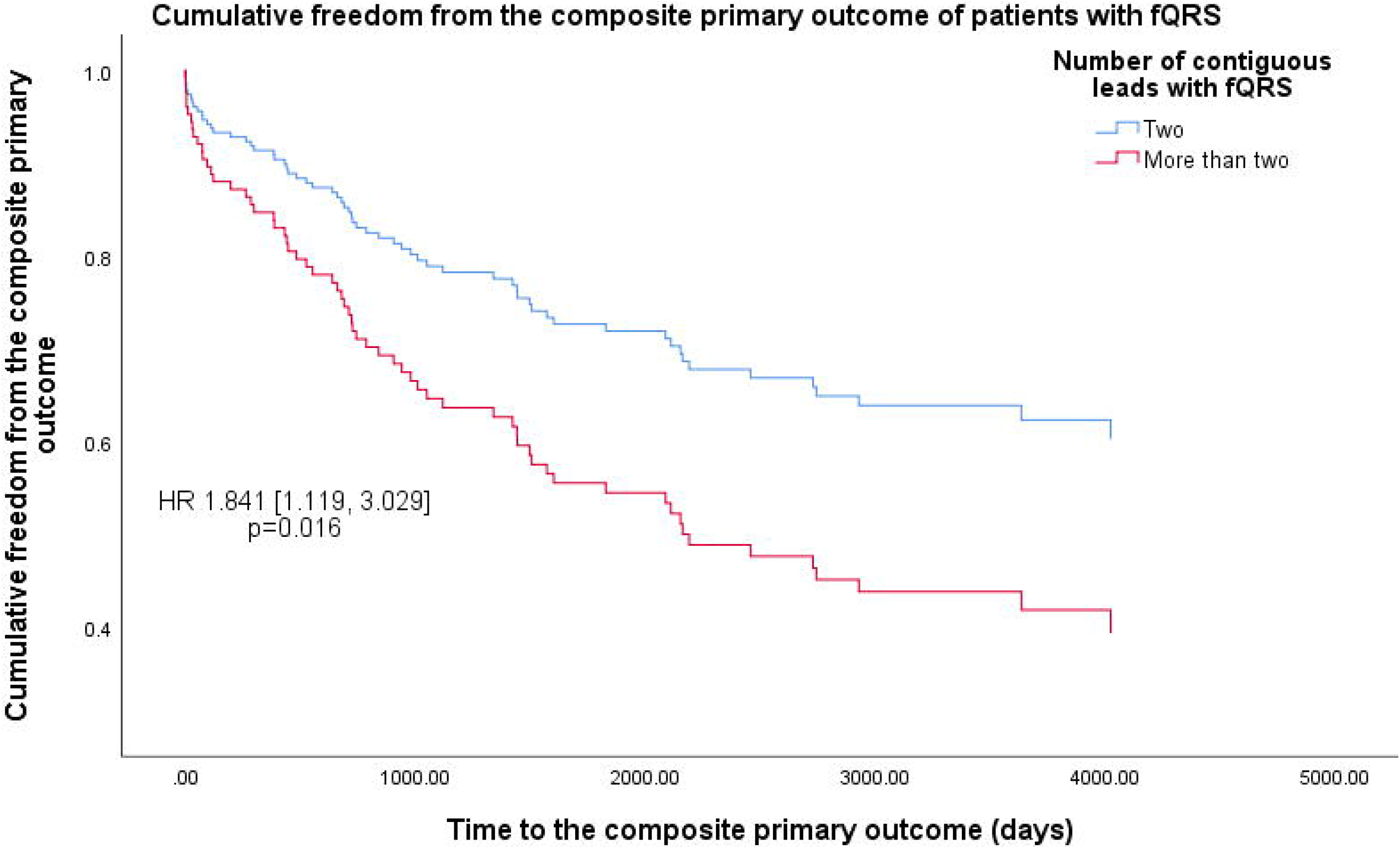
Kaplan-Meier curves of cumulative freedom from the primary composite outcome of patients with fragmented QRS (fQRS), stratified by fQRS burden. The hazard ratio (HR) shown was adjusted for differences in baseline characteristics.

## 4. Discussion

In this retrospective cohort study of heart failure patients, fQRS is strongly and independently predictive of cardiovascular mortality, VA and SCD, with the presence of fQRS in more than two contiguous leads being associated with significantly higher risk of SCD. Additionally, we showed that while fQRS was strongly predictive of VA and SCD regardless of IHD status, though fQRS was only predictive of cardiovascular mortality in patients with IHD.

Previous studies have shown that fQRS is predictive of mortality and arrhythmic events in patients with heart failure.(4) To the authors’ best knowledge, the present study is the first study focusing on Asian patients hospitalized with heart failure that reported long-term clinical outcomes, with a mean follow-up duration exceeding five years. We confirmed that the elevated risk of adverse outcomes in patients with fQRS persists in the long term, independent of prior history of VA. The magnitudes of elevated risk we observed were also comparable to previous studies.(4) Previous studies on Asian patients with heart failure have found fQRS in 16.6-49% of patients.(6,10,11) In contrast, we identified fQRS in 8.20% of included patients. This difference in prevalence was probably due to the less-selective inclusion and exclusion criteria we used. Having identified all patients through a territory-wide database, the results of our study more closely reflect real life practice, and better guide clinicians in their care of patients with heart failure.

Additionally, we found that a higher burden of fQRS was associated with further increased risk of SCD. A similar concept has been shown by Debonnaire et al in patients with hypertrophic cardiomyopathy.(12) This finding may be explained mechanistically by higher fQRS burden reflecting more extensive myocardial scarring.(13) By choosing a cutoff of two contiguous leads with fQRS, our results meant that the presence of additional leads with QRS complexes of fragmented morphology beyond the classification criteria would be additionally predictive of SCD. This allowed for a straightforward and intuitive interpretation, potentially facilitating clinical applications.

Importantly and interestingly, we showed that fQRS was predictive of SCD but not cardiovascular mortality in patients without IHD. It must be noted that non-ischaemic cardiomyopathies are a heterogeneous entity with various aetiologies, and the pathophysiological changes leading to the presence of fQRS might vary accordingly. It is thus possible that fQRS has varying significance in these patients depending on the aetiology of cardiomyopathy. Further studies are warranted in this area.

### 4.1. Limitations

This study has several limitations. First, data obtained from CDARS could not be adjudicated. However, all data entry were done by clinicians who were not involved in this study. Second, without accompanying echocardiographic and aetiological data, the included patients with heart failure were heterogeneous in aetiology and phenotype. As shown by the subgroup analysis on patients with or without IHD, such differences may affect the prognostic value of fQRS. Third, our analysis did not account for differences in the morphologies of fQRS, which may have prognostic implications.(2)

## 5. Conclusion

The presence of fQRS was independently predictive of cardiovascular mortality, VA, and SCD in Asian patients hospitalized for heart failure. Having fQRS in more than two contiguous leads independently predicted further increased risk of SCD. However, fQRS did not predict cardiovascular mortality in patients without IHD, warranting further studies.

## Data Availability

The minimal dataset underlying this study is available upon reasonable request to the corresponding author.

## 10. Supplementary materials

Supplementary Table S1. Cox regression results, stratified by the presence of ischaemic heart disease. Hazard ratios (HR) were referenced against patients without fragmented QRS.

Supplementary Table S2. Cox regression of the 179 patients with fragmented QRS. Hazard ratios were referenced against patients with fragmented QRS present in only two contiguous leads.

