## Supplementary Table for "Fragmented QRS is independently predictive of long-term adverse clinical outcomes in Asian patients hospitalized for heart failure: a retrospective cohort study"

**SUPPLEMENTAL MATERIAL**

**Supplementary Table S1.** Cox regression results, stratified by the presence of ischaemic heart disease. Hazard ratios (HR) were referenced against patients without fragmented QRS.

| Subgroup | Outcome | | Univariate | | Multivariate[1] | |
|---|---|---|---|---|---|---|
| | | | HR [95% CI] | p value | HR [95% CI] | p value |
| Ischaemic heart disease present (N=786) | Composite primary outcome[2] | | 1.703 [1.166, 2.488] | 0.006 | 1.746 [1.191, 2.559] | 0.004 |
| | Secondary outcomes | Cardiovascular mortality | 1.745 [1.142, 2.668] | 0.010 | 1.802 [1.173, 2.768] | 0.007 |
| | | Ventricular arrhythmia | 2.842 [1.152, 7.014] | 0.023 | 2.652 [1.065, 6.602] | 0.036 |
| | | Sudden cardiac death | 1.831 [0.993, 3.376] | 0.053 | 1.881 [1.014, 3.481] | 0.045 |
| | | Myocardial infarction | 1.085 [0.684, 1.722] | 0.728 | 1.052 [0.661, 1.672] | 0.832 |
| | | New-onset atrial fibrillation | 1.072 [0.709, 1.620] | 0.742 | 1.116 [0.737, 1.692] | 0.604 |
| No ischaemic heart disease (N=1396) | Composite primary outcome[2] | | 1.334 [0.926, 1.920] | 0.121 | 1.342 [0.929, 1.938] | 0.117 |
| | Secondary outcomes | Cardiovascular mortality | 1.062 [0.672, 1.678] | 0.797 | 1.049 [0.662, 1.662] | 0.839 |
| | | Ventricular arrhythmia | 3.487 [1.414, 8.601] | 0.007 | 3.336 [1.343, 8.282] | 0.009 |
| | | Sudden cardiac death | 1.809 [1.071, 3.053] | 0.027 | 1.928 [1.135, 3.278] | 0.015 |
| | | Myocardial infarction | 0.788 [0.402, 1.547] | 0.489 | 0.895 [0.454, 1.762[ | 0.747 |
| | | New-onset atrial fibrillation | 1.168 [0.870, 1.570] | 0.302 | 0.835 [0.620, 1.125] | 0.235 |

[1] Adjusted for hypertension for patients with ischaemic heart disease, and renal diseases, atrial fibrillation, diabetes mellitus, calcium channel blocker usage, and usage of statins and fibrates for patients without ischaemic heart disease

[2] A composite of cardiovascular mortality, ventricular arrhythmia, and sudden cardiac death

CI, confidence interval.

**Supplementary Table S2.** Cox regression of the 179 patients with fragmented QRS. Hazard ratios were referenced against patients with fragmented QRS present in only two contiguous leads.

| Outcome | | Hazard ratio [95% confidence interval]) | p value |
|---|---|---|---|
| Composite primary outcome[1] | | 1.841 [1.119, 3.029] | 0.016 |
| Secondary outcomes | Cardiovascular mortality | 1.539 [0.850, 2.786] | 0.155 |
| | Ventricular arrhythmia | 1.897 [0.611, 5.888] | 0.268 |
| | Sudden cardiac death | 2.866 [1.350, 6.081] | 0.006 |
| | Myocardial infarction | 1.746 [0.835, 3.649] | 0.139 |
| | New-onset atrial fibrillation | 1.096 [0.676, 1.775] | 0.710 |

[1] A composite of cardiovascular mortality, ventricular arrhythmia, and sudden cardiac death
